# Is Wear Still a Concern in TKA With Contemporary Conventional and Highly Crosslinked Polyethylene Tibial Inserts in the Mid- to Long-Term?

**DOI:** 10.1101/2024.05.29.24308156

**Authors:** Devin P. Asher, Jennifer L. Wright, Deborah J. Hall, Hannah J. Lundberg, Douglas W. Van Citters, Joshua J. Jacobs, Brett R. Levine, Robin Pourzal

**Author notes:** **Corresponding Author:** Robin Pourzal, PhD, 1611 W. Harrison St. Ste. 204-F Chicago, IL 60612.

## Abstract

**Background:** Modern literature has brought into question if wear of tibial inserts made from conventional or highly-crosslinked polyethylene (HXL PE) is still a factor limiting longevity of total knee arthroplasty (TKA) in the mid- to long-term. It is the objective of this study to determine: 1) most common causes of mid- to long-term TKA failure, 2) the prevalence of delamination, and 3) the medial/lateral linear wear rates of conventional and HXL PE tibial inserts retrieved in the mid- to long-term.

**Methods:** A tibial insert retrieval cohort of 107 inserts (79 conventional, 28 HXL PE) with a minimum time *in situ* of 6.5 years (mean 11.7±4) was studied. Failure causes were determined from chart-review, delamination presence was assessed microscopically, and medial/lateral linear wear was determined by minimal thickness changes measured with a dial-indicator.

**Results:** The most common mid-to long-term etiologies for failure were instability (44.9%), PE wear 15%), aseptic loosening (14%) and infection (13.1%). Delamination occurred in 70% of inserts (72.1% conventional, 64.3% HXLPE). Gross material loss due to delamination appeared to be the underlying reason for at least 33.3% of cases exhibiting instability. Of the cases removed for infection, 75% exhibited no histopathological hallmarks of acute infection. The medial/lateral wear rates were 0.054/0.051 (conventional) and 0.014/0.011 (HXL) mm/year, respectively.

**Conclusions:** Polyethylene wear still appears to be a major primary and secondary cause for TKA revision in the mid- to long-term. Wear may manifest as destabilizing delamination or as continuous release of fine wear particles potentially resulting in inflammatory responses and subsequent failure.

## Introduction

Total knee arthroplasty (TKA) is performed on more than one million patients in the US annually to treat end-stage osteoarthritis (OA). Revision rates, however, have remained steady in recent years. (1) According to registries and epidemiological studies, the most common causes of failure leading to revision include infection and inflammatory responses, mechanical loosening, and instability, with the proportion of revisions due to aseptic loosening decreasing in favor of infection. (2–5) One limitation of registry reports is that they aggregate early and late failures, which typically represent different rates of etiologies for revision. Therefore, as newer registries increase reporting, they likely overrepresent early failures predominately related to higher rates of early prosthetic joint infection (PJI). (4)

Historically, two factors limited long-term implant survivorship: shelf and *in vivo* ultra-high molecular weight polyethylene (PE) oxidation leading to mechanical insert breakdown and particle-induced osteolysis driven by PE wear debris leading to subsequent implant loosening. The advent of highly crosslinked (HXL) PE has led to decreases in revision rates due to wear in the short term (6,7), but long-term clinical and retrieval studies of contemporary HXL PE have not yet been published, particularly with respect to *in vivo* oxidation prevention and long-term biological outcomes. One registry found improved mid-term survivorship (9), but most studies find no difference between conventional and HXL PE performance for TKA in terms of revision rate, cause for revision, or patient satisfaction. (2,6–8) Retrieval studies of contemporary tibial inserts—made from either PE—retrieved at revision surgery indicate that *in vivo* oxidation is frequent, even in the absence of free radicals. (9–11)

The current literature reports varying revision rates for wear, with two studies finding unchanged wear-related failures over time. (4,12) Confounding the issue is the difficulty of stratifying reasons for aseptic revisions within registries, based on overall coding discrepancies and lack of transparency. (13) Furthermore, PE wear, especially gross wear caused by delamination, may be the underlying cause of the physical examination finding of instability. However, PE wear may not be included in the coding process as it is often an intra-operative finding not discernable on preoperative evaluation.

Moreover, the AJRR reports that only 3% of failures today are associated with wear particle-induced osteolysis. However, with nearly half of all revision cases being attributed to aseptic loosening and non-infectious inflammatory reasons, it is possible that biological reactions to particulate debris still occur on a longer timeline. Further, the reaction to particulate debris may be a secondary factor in a significant number of revision procedures and likely is highly underreported.

Therefore, this study aims to answer the following research questions: 1) What are the most common causes of mid- to long-term TKA failure with conventional PE and contemporary HXL PE inserts? 2) What is the prevalence of delamination, and how does it relate to the cause of failure? 3) What are the medial and lateral linear wear rates of conventional PE and contemporary HXL PE inserts measured from mid- to long-term retrievals?

## Material and Methods

A retrospective chart review was conducted to determine general information on all surgically retrieved TKAs made from conventional PE (retrieved between 2019 and 2024) and HXL PE (retrieved between 2014 and 2024) available in our IRB-approved implant retrieval repository. Data collected included time *in situ*, sex, surgery site laterality, and reason for implant/removal, as well as implant manufacturer, model, and size. Inserts made from conventional or HXL PE with a time *in situ* of > 6.5 years, representing mid- to long-term implants, were included in this study. After performing a review of 688 patients, a total of 110 tibial inserts fit the criteria for the study. Three inserts made from antioxidant doped polyethylene were removed from the study resulting in a final study cohort of 107 inserts. The mean (standard deviation, SD) patient age at revision was 71 (SD 7) years, the mean time in situ was 12 (SD 4) years. There were 63 female and 44 male patients (Table 1). This cohort included 79 conventional and 28 HXL PE inserts from 6 manufacturers (71 Zimmer Biomet—Warsaw, IN, 8 Smith & Nephew—Memphis, TN, 2 DePuy—Warsaw, IN, 18 Stryker—Mahwah, NJ, 5 Biomet—Warsaw, IN, 3 other). There were 81 cruciate retaining inserts and 26 posterior stabilized inserts.

**Table 1.** Demographics of the study cohort.

|  | Age (years) | Time in situ (years) | Sex (female, male) | Liner type (cruciate retaining or posterior stabilized) |
| --- | --- | --- | --- | --- |
| All | 71.16 ± 6.72 | 11.68 ± 3.94 | 63, 44 | 81 CR, 26 PS |
| Conventional PE | 70.55 ± 6.71 | 12.49 ± 4.01 | 46, 33 | 67, 12 |
| HXL PE | 72.90 ± 6.58 | 9.40 ± 2.71 | 17, 11 | 14, 14 |

**Table 2.** Medial and lateral linear wear and wear rate for inserts made from conventional and HXL PE.

|  | medial linear wear (mm) | lateral liner wear (mm) | medial linear wear without delamination (mm) | lateral liner wear without delamination (mm) | medial linear wear rate (mm/year) | lateral liner wear rate (mm/year) |
| --- | --- | --- | --- | --- | --- | --- |
| Conventional PE | 0.78<br>(0.09, 4.26) | 0.7<br>(0, 2.1) | 0.69<br>(0.09, 2.69) | 0.67<br>(0, 1.44) | 0.054<br>(0, 0.17) | 0.051<br>(0, 0.12) |
| HXL PE | 0.12<br>(0, 2.6) | 0.093<br>(0, 1.65) | 0.11<br>(0, 0.54) | 0.084<br>(0, 0.7) | 0.014<br>(0, 0.07) | 0.011<br>(0, 0.05) |

Tibial insert wear and damage was first optically assessed under a stereo microscope. A delamination score was assigned for the medial and lateral sides using a three-tiered numeric rating system where 0 = no visible delamination is present, 1= mild (visible subsurface cracks, milky appearance), and 2 = severe (gross wear and breakdown of the bearing surface) (Figure 1).

**Figure 1.**
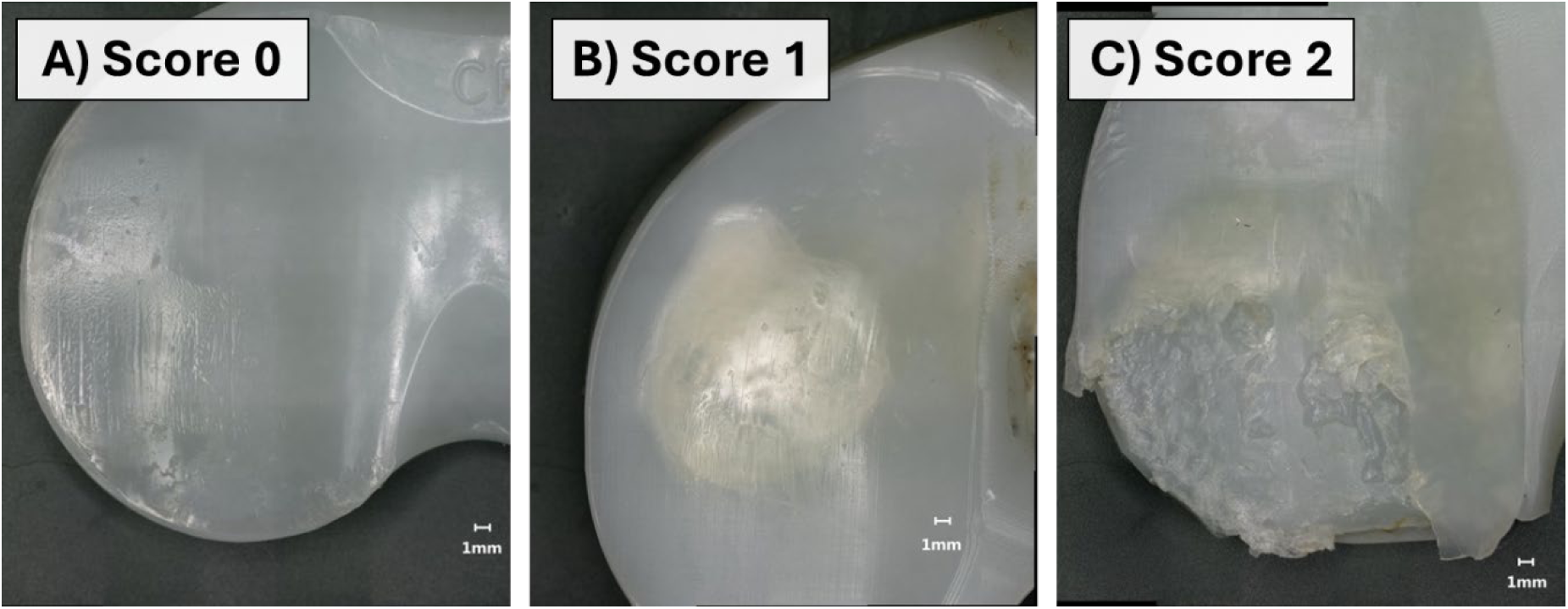
Photographs illustrating characteristic features of various degrees of delamination: A) The PE bearing surface exhibits characteristic wear features such as polishing, scratches, striated patterns, and pitting, however, no delamination was noted. B) Subsurface cracks can be seen on the bearing surface characterized by a ‘milky’ surface appearance, but there is no significant material loss. C) Gross material loss associated with delamination on the bearing surface. Note that delamination scores 1 and 2 can vary in the number of affected locations and overall area.

Linear wear was quantified according to a previously published method based on change of minimum thickness (14,15) using a Mitutoyo dial indicator. A 3mm spherical tip was used on both sides of the indicator (Figure 2). Each insert was evaluated for linear wear on the medial and lateral articular surfaces at the narrowest point of the bearing surface. This process was repeated for a total of three measurements per side. Linear wear on each side was defined as the difference between the measured value and the initial minimum thickness. Reference values for the minimal thickness were obtained from either publicly available product information or from short-term retrievals (time *in situ* < 6 months). Linear wear was measured for all inserts, even those with severe delamination, to determine the maximum linear wear independent of damage mechanism. However, because delamination is an erratic process that does not necessarily correlate with the release of wear particles, linear wear rates were only assessed for inserts that were not severely delaminated (i.e. those with bearing surfaces undergoing sliding wear without gross wear due to delamination).

**Figure 2.**
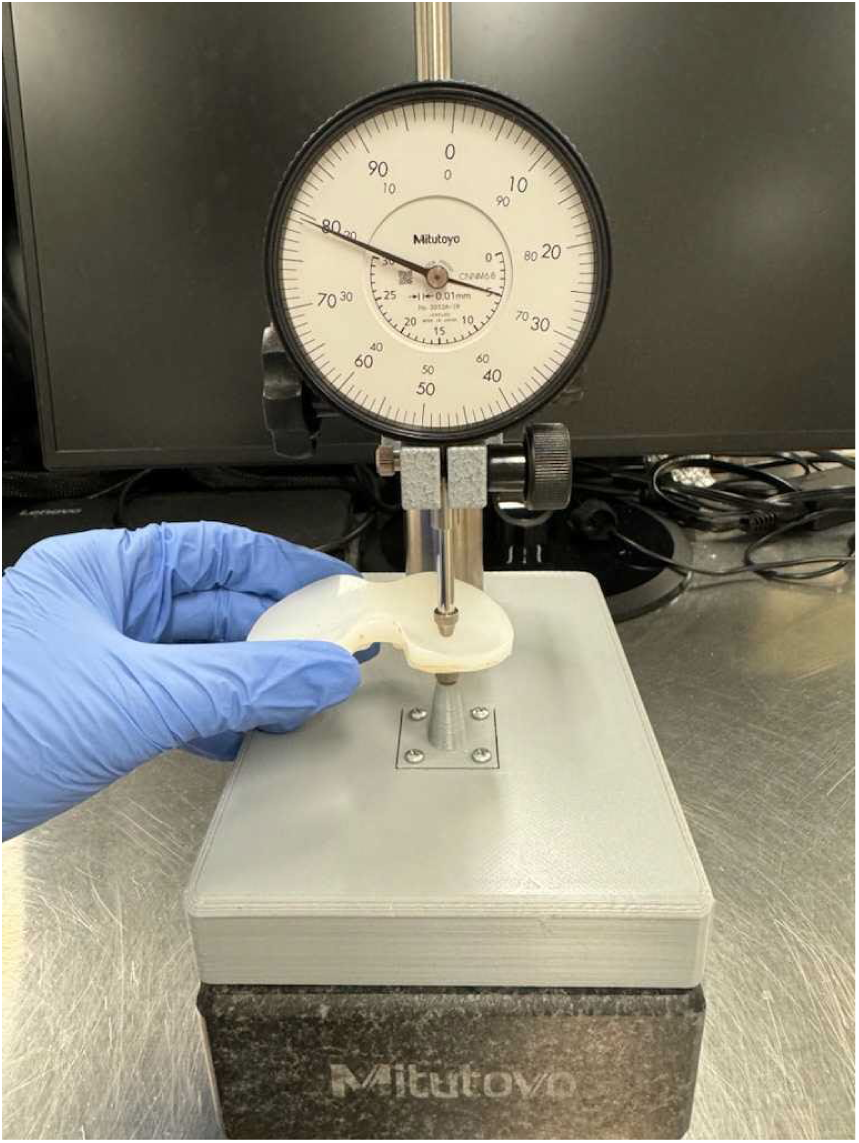
Measuring rig used for thickness change evaluation as a proxy measure for linear wear.

## Results

In this tibial insert retrieval cohort, the primary reasons for revision were 44.9% instability (48.1% conventional PE, 35.7% HXL PE), 15% polyethylene wear (16.5% conventional PE, 10.7% HXL PE), 14% aseptic loosening (13.9% conventional PE, 14.3% HXL PE), and 12.1% infection (13.9% conventional PE, 7.1% HXL PE) (Figure 3). The other reasons for revision included 4.7% pain (2.5% conventional PE, 10.7% HXL PE), 3.7% extensor mechanism disruption (1.3% conventional PE, 10.7% HXL PE), 1.9% osteolysis (1.3% conventional PE, 3.6% HXL PE), 2.8% stiffness (1.3% conventional PE, 7.1% HXL PE), and 0.9% distal femur fracture (1.3% conventional PE, 0% HXL PE). With respect to the PJI cases, review of the pathology reports revealed that only 2 of 8 (25%) cases exhibited acute infection, while the others were classified as inflammatory reactions.

**Figure 3.**
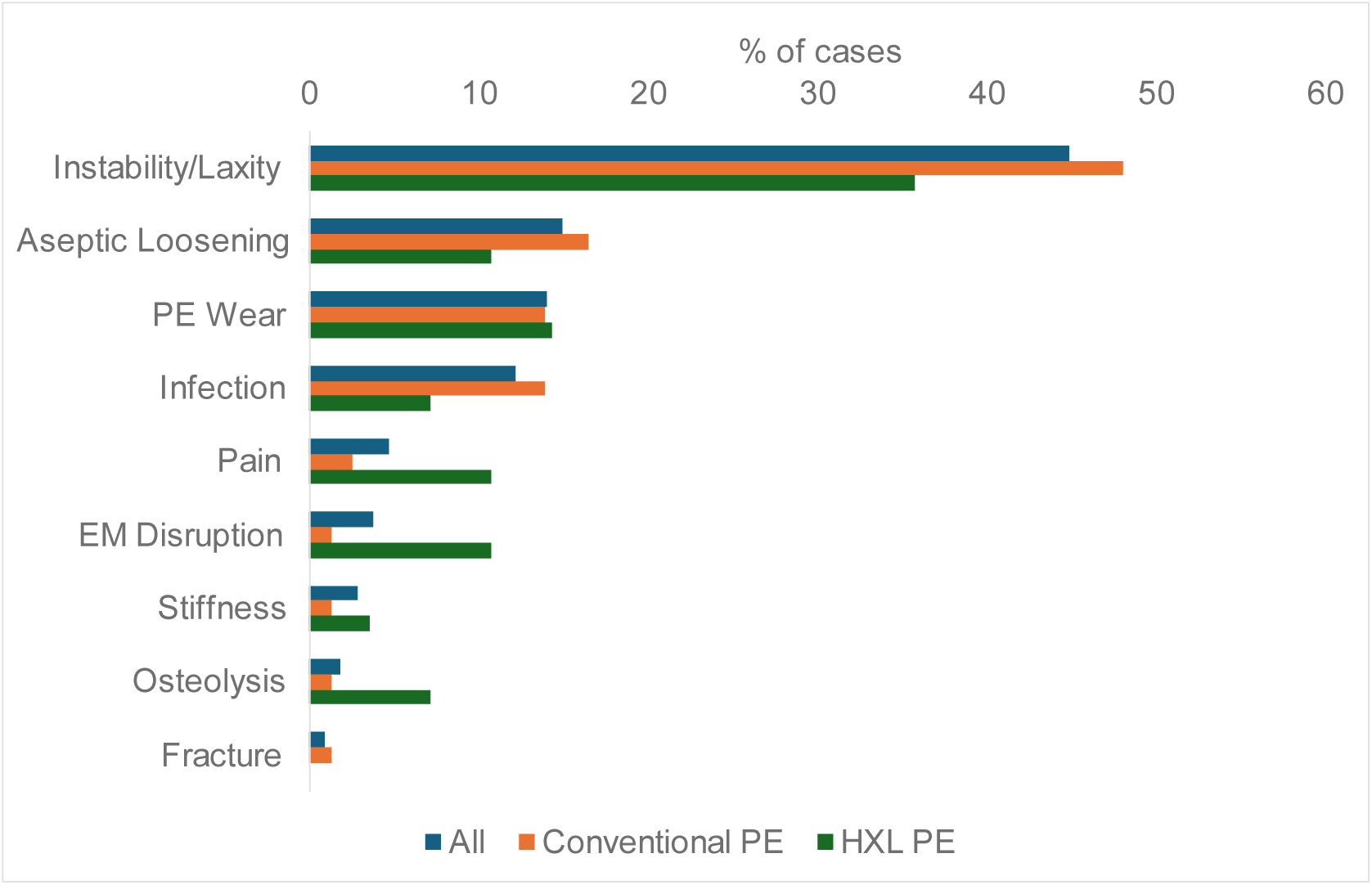
Illustration of the main reason for mid- to long-term failures associated with this tibial insert cohort and breakdown by type of polyethylene.

Delamination occurred in 70% of inserts with 36.4% exhibiting mild and 33.6% severe delamination. Both conventional and HXL PE inserts had delamination (72.1% conventional PE: 32.9% mild,39.2% severe; 64.3% HXL PE: 46.4% mild, 17.9% severe). If delamination was present, it occurred on both the medial and lateral tibial condyles in 54.7% (conventional PE 56.1%, HXL PE 50%) of cases, only the medial condyle in 32% (conventional PE 33.3%, HXL PE 27.8%), only the lateral condyle in 9.3% (conventional PE 8.8%, HXL PE 11.1%), and only on the central post in 4% (conventional PE 1.8%, HXL PE 11.1%).

When stratified by reason for revision, delamination occurred in 68.7% (35.4% mild, 33.3% severe) of inserts with instability, 93.8% (31.3% mild, 62.5% severe) of inserts removed for wear, 86.7% (46.7% mild, 40% severe) of inserts that underwent aseptic loosening, and 53.9% (30.8% mild, 23.1% severe) of insert removed for infection or other inflammatory responses (Figure 4).

**Figure 4.**
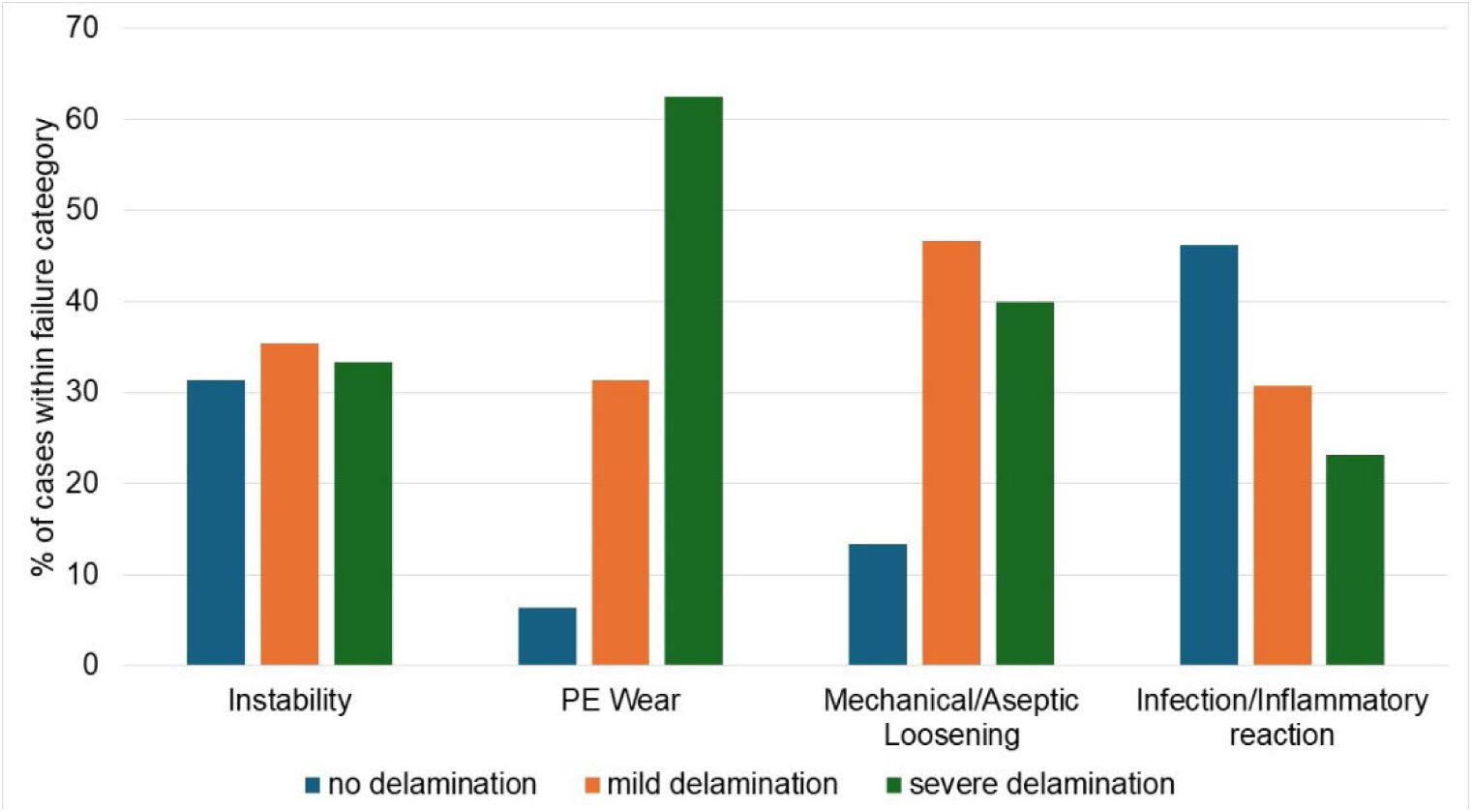
Distribution of delamination for tibial inserts grouped by the major reasons for failure in this cohort.

Linear wear could be assessed for all inserts except of six (all conventional PE), for which no reference was available. The medial and lateral median (minimum, maximum) linear wear was 0.78 mm (0.09, 4.26) and 0.7 mm (0, 2.1) for conventional PE, and 0.12 mm (0, 2.6) and 0.11 mm (0, 1.65) for HXL PE inserts, respectively. When excluding inserts with severe delamination, the medial and lateral median (min., max.) linear wear was 0.69 mm (0.09, 2.69) and 0.67 mm (0, 1.44) for conventional PE, and 0.11 mm (0, 0.54) and 0.084 mm (0, 0.7) for HXL PE inserts, respectively. Including inserts with severe delamination there was no linear relationship between wear and time *in situ* for either PE type, except for a positive correlation on the lateral side of conventional PE inserts (R^2^=0.14.5, p=0.001). Excluding inserts with severe delamination, conventional PE inserts had a positive correlation between linear wear and time *in situ* on both the medial (R^2^=0.11.6, p=0.008) and lateral side (R^2^=0.20, p<0.001). For HXL PE inserts without severe delamination, only the lateral side exhibited a positive correlation between wear and time *in situ* (R^2^=0.30, p=0.006) (Figure 5).

**Figure 5.**
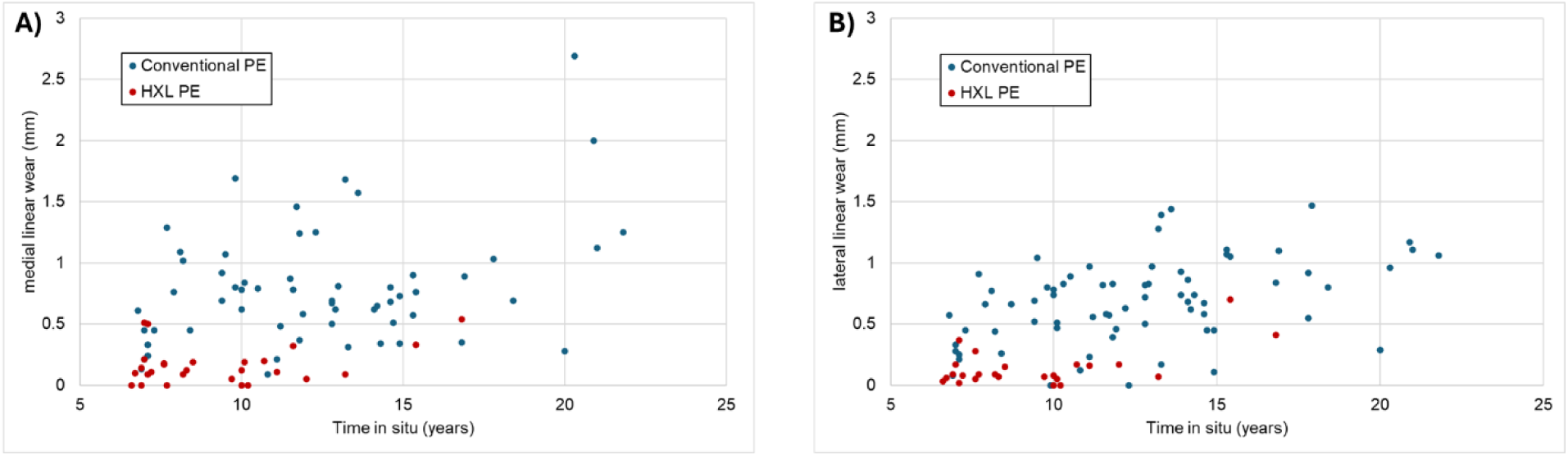
Scatter plots illustrating linear wear over time *in situ* on the A) medial, and B) lateral side.

Wear rates were computed for inserts without severe delamination as the median linear wear over time *in situ*. The median medial and lateral wear rates for conventional and HXL PE were 0.054 mm/year (0, 0.17), 0.051 mm/year (0, 0.12), 0.014 mm/year (0, 0.07), and 0.011 mm/year (0, 0.05), respectively.

## Discussion

Instability, PE wear, aseptic loosening and inflammatory responses were the most common mid- to long-term failure reasons for all retrievals whether made of conventional or HXL PE. Delamination and progressive surface wear were also prevalent findings within all reasons for revision. This is a departure from recent registry reports which found infection and other inflammatory responses to be the most common cause of failure. (2) However, registry data is skewed by early failures and therefore are not a reliable measure for mid- to long-term revision causes.

The most dominant mid- to long-term TKA failure reason was instability, which coincided with delamination in 68.7% of cases, of which 33.3% was severe delamination damage with material loss. Instability is commonly attributed to malpositioning, cement debonding, or other mechanical issues, which would likely manifest within the first 5 years after primary surgery. (16–21) Therefore, it is plausible that instability in the mid- to long-term frequently may occur secondary to delamination. Delamination, which is the result of surface fatigue wear under combined rolling and sliding motion in the knee, has been a prominent problem in TKA with tibial inserts made from historic PE leading to early failures. (22,23) Embrittlement of the material due to *in vivo* oxidation enables this wear mechanism. Improvements to the material by vitue of sterilization and packaging in an inert atmosphere and the introduction of highly crosslinked materials have mitigated this issue. (24,25) However, several studies have demonstrated that contemporary conventional and HXL PE inserts also undergo subsurface *in vivo* oxidation although on a longer time scale. (10,26,27) The maximum oxidation index is typically located a few hundred micrometers underneath the bearing surface. Our study demonstrates that delamination persists in contemporary conventional and HXL PEs for tibial inserts retrieved in the mid- to long-term. Gross wear associated with delamination may cause instability of the joint or accelerate the process from sub-clinical to symptomatic instability. Even the presence of multiple subsurface cracks that have not yet caused material breakdown may change the mechanical and wear behavior of the insert. While the problem of *in vivo* oxidation has been further addressed by the introduction of antioxidant doped polyethylene (6,7,28), our repository did not yet have a sufficient number of retrievals with the required implantation duration available to investigate trends with these newer materials.

After instability, the main reasons for revision were PE wear, aseptic loosening, and PJI or other inflammatory responses. PE wear cases all exhibited some degree of delamination, which was not surprising because delamination is characterized by severe damage to the articular surface. Such gross PE wear can often be assessed as eccentric wear on preoperative radiographs. In the American Joint Replacement Registry (AJRR) PE wear and osteolysis cases are grouped together as a single category, based on ICD 10 coding parameters. In the current study, the two failure reasons were separated because the underlying damage mechanisms are different. Osteolysis presents with obvious osteolytic lesions/bony changes on preoperative radiographs (29), while PE wear cases undergo delamination with material loss most commonly identified intraoperatively. Osteolysis is frequently driven by a foreign body particle response to small particles on the scale of tens to hundreds of nanometers (29–31). Particles trigger a macrophage response that can initiate a cascade of cell responses resulting in periprosthetic bone loss and loosening. Delamination leads to the release of larger particles which may also trigger a tissue response, especially the formation of foreign body giant cells, but not necessarily osteolysis (32,32). While inserts undergoing delamination also generate fine particles during articulation, there is also concern of mechanical failure besides potential inflammatory reactions..

Particle induced osteolysis or inflammatory response may also be responsible for failure cases categorized as aseptic loosening and infection. As stated above, it can be assumed that the majority of aseptic loosening cases are not related to mechanical issues such as malpositioning or cement debonding as those would likely manifest early. (16–21) Thus, in the current study of mid- to long-term failures, early stage osteolysis caused by wear debris or other foreign bodies (e.g. cement debris, corrosion products, etc.) may be a possible reason for failure. Additionally, 75% of cases labeled as infection related failures did not exhibit hallmarks of PJI according to pathology reports, suggesting a likely inflammatory response to wear debris.

The wear analysis in this study clearly shows that measurable wear occurs during the normal articulation of the knee for both conventional and HXL PE. While there was no or only a weak correlation between wear and time *in situ*, it must be considered that there are multiple other factors that drive wear (sex, implant positioning, design, etc.) which could not be assessed here due to the relatively small numbers involved after stratification for these factors. (33) It was of note that the linear wear rates for this mid- to long-term retrieval study were similar to previous studies using the same dial method, (9) but lower than that reported by surface reconstruction methods (33). Yet, it is of note that the latter study focused on early to mid-term TKA failures of a single design. Both the wear rate and the incidence of delamination were higher on the medial compared to lateral compartment, which was expected based on the higher joint loading and surface pressure on the medial side. Joint loading on the medial side has been reported as up to 90% of the total force through the joint depending on TKA design due to knee alignment and to the action of the abductors during *in vivo* activity. (34,35)

It is important to state that the majority of particles generated from contemporary PE inserts, especially HXL PE inserts, are too small to be identified using polarized light microscopy. However, recent studies of HXL PE acetabular inserts in total hip replacement have shown a prominent presence of PE debris within periprosthetic macrophages by means of infrared spectroscopic imaging.(36,37) In fact, it was shown that osteolysis could occur in inserts with a total volumetric wear rate comparable to well-functioning inserts made from historic PE. These findings demonstrate that despite less volumetric wear in HXL PE inserts, the finer particles generated from HXL PE carry a larger osteolytic potential. (38,39) Future studies need to examine the tissue response of mid- to long-term TKA in more detail as well as the possible association of failure with intracellular fine polyethylene debris.

This study had several limitations. First, the study cohort is too limited in size to differentiate differences between tibial insert designs and manufacturers, different insert sizes, or between cruciate retaining and posterior stabilized inserts. It is of great importance to increase this cohort of mid- to long-term retrievals to gain a better understanding of the *in vivo* wear performance of contemporary TKA. Second, an important comparison group—antioxidant doped inserts—could not be included because too few were available in our retrieval database at this time. Third, this study was limited to linear wear measurements based on thickness change. Yet, linear wear was shown to be a good surrogate measure for wear to accurately reflect trends in wear behavior of a retrieval cohort, while underestimating wear on an individual basis. (40) Another limitation of this study is the differences among both conventional and HXL PEs across manufacturers. While treated as only two different groups in this study, the degree of crosslinking and resulting mechanical properties are likely to differ among HXL inserts resulting in additional confounders. (9,14) Finally, this study lacked histopathological analysis of corresponding periprosthetic tissues because tissue samples were not available for all cases. Future studies need to investigate the relationship between wear volume, extent of macrophage response, particle size, and the ultimate reason of failure.

## Conclusions

This study demonstrates that the reasons for TKA failure in the mid- to long-term differ substantially from registry studies that include early failure. While infection and other inflammatory responses are the most common reason for revision according to the AJRR, in this retrieval cohort the predominant reason for failure was instability. Only 8 of 107 inserts failed due to infection and other inflammatory reactions in our cohort, and only two of these 8 demonstrated histopathological findings consistent with periprosthetic joint infection. In both conventional and HXL PE tibial inserts, implant wear can still manifest as 1) delamination leading to gross wear, and 2) milder wear—usually characterized by polishing and three-body wear—resulting in the continuous generation of fine wear debris. The results of this study suggest that delamination may be linked to the onset of instability in the mid- to long-term due to the breakdown of the bearing surface and loss of structural integrity. Other than instability, tibial inserts were removed predominantly for aseptic loosening, PE wear, and other inflammatory responses. The diagnosis of PE wear related TKA failure was mostly linked to the occurrence of delamination. Considering that wear rates were measurable for both conventional and HXL PE, and the fact that loosening due to mechanical issues commonly manifest early, it is likely that the accumulation of wear debris within periprosthetic tissue can cause an inflammatory response resulting in aseptic loosening, effusion, and pain. While only 2% of cases in this cohort were diagnosed with osteolysis, it appears likely that this number is underestimated. The success of HXL PE in total hip arthroplasty may have resulted in a false sense of security for total knee arthroplasty, and only ongoing long-term studies will tell us if PE wear-related issues are truly eliminated. This study suggests that PE wear resulting in loss of structural integrity, manifesting as instability, and/or inflammatory tissue response to wear debris is still a major reason for TKA failure in the mid- to long-term.

## Data Availability

All data produced in the present study are available upon reasonable request to the authors.

## Acknowledgements

This study was funded by an internal pilot grant from Rush University Medical Center (Rush Catalyst Award, PIs Lundberg and Pourzal). The authors would like to thank Julia Hochstatter for her valuable contributions in the laboratory.

